# An optimized MRI and PET based clinical protocol for improving the differential diagnosis of Geriatric Depression and Alzheimer’s Disease

**DOI:** 10.1101/2021.02.08.21251205

**Authors:** Louise Emsell, Heleen Vanhaute, Kristof Vansteelandt, François-Laurent De Winter, Danny Christiaens, Jan Van den Stock, Rik Vandenberghe, Koen Van Laere, Stefan Sunaert, Filip Bouckaert, Mathieu Vandenbulcke

## Abstract

**OBJECTIVE:** MRI derived hippocampal volume (HV) and amyloid PET may be useful clinical biomarkers for differentiating between geriatric depression and Alzheimer’s Disease (AD). Here we investigated the incremental value of HV and 18F-flutemetmol PET in tandem and sequentially to improve discrimination in unclassified participants.

**METHOD:** Two approaches were compared in 41 participants with geriatric depression and 27 participants with probable AD: (1) amyloid and HV combined in one model and (2) HV first and then amyloid.

**RESULTS:** Both HV(χ^2^(1) = 6.46: p= 0.011) and amyloid (χ^2^(1) =11.03: p=0.0009) were significant diagnostic predictors of depression (sensitivity: 95%, specificity: 89%). (2) 51% of participants were correctly classified according to clinical diagnosis based on HV alone, increasing to 87% when adding amyloid data (sensitivity: 94%, specificity: 78%).

**CONCLUSION:** Hippocampal volume may be a useful gatekeeper for identifying depressed individuals at risk for AD who would benefit from additional amyloid biomarkers when available.

## Introduction

Alzheimer’s disease (AD) and geriatric depression are two of the most prevalent disorders in the elderly. Differential diagnosis between these conditions is challenging in clinical practice given their shared psychopathology (1-3). For example, in contrast to early-onset cases of depression (<60 years old), people with geriatric depression are more likely to present with cognitive changes such as impaired memory function, attentional difficulties and executive dysfunction. These symptoms may overlap with the first clinical presentation of prodromal or early AD. On the other hand, depressive symptoms are frequently seen in mild cognitive impairment (MCI) and AD (4-6). Increasing evidence linking depression with subsequent cognitive decline makes diagnosis even more challenging. The most important hypotheses which address this association are the stress-hypothesis, which proposes that prior experience of major depressive disorder (MDD) is a risk factor for developing AD as a result of stress-mediated physiological effects that accelerate brain ageing (7-9) and the neuropsychiatric hypothesis, which views MDD as a prodrome of AD and therefore presupposes common pathophysiological mechanisms (10-12). In line with this last hypothesis, the concept of ‘mild behavioral impairment’ (MBI) has been constructed to describe sustained and impactful late life behavioral changes, such as affective dysregulation or decreased motivation, that cannot be better attributed to a psychiatric or medical disorder (13). Similar to people with MCI, those with MBI are at risk for cognitive decline and all-cause dementia. Changes in MBI subdomains of motivation or affect may predetermine evolution to AD (14, 15).

Owing to the challenges in differentiating geriatric depression from AD, misdiagnosis is frequent with 15-23% of people with AD receiving a prior psychiatric diagnosis and, conversely, around 30% of people with geriatric depression being erroneously diagnosed with AD (13, 16). Yet, a timely and correct diagnosis is critical for the initiation of a suitable treatment (which could possibly reverse cognitive deficits in depression), avoiding unnecessary diagnostic tests and, in the case of AD, developing future interventions.

Given that clinical symptoms are often insufficient to make an accurate diagnosis, there is an increasing need for reliable in vivo biomarkers. MRI-based hippocampal atrophy is one of the biomarkers used as a supporting feature in several revised diagnostic criteria for mild cognitive impairment (MCI) and AD as an indicator for neuronal injury (N) in the ATN (amyloid-tau-neuronal injury) model of the AD continuum (17-19). At the same time, hippocampal volume loss has been frequently associated with geriatric depression (20-23). Amyloid imaging using PET is another important biomarker in diagnosing AD assessing the A-status of patients and has a high negative predictive value. Especially in the elderly population, age-dependent increased amyloid deposition without cognitive impairment occurs (24) decreasing the specificity of amyloid PET with advanced age. Amyloid imaging can increase diagnostic accuracy particularly in people with possible AD presenting with unclear clinical presentation, atypical clinical course or etiologically mixed presentation (25). However, in general it is currently less widely available compared to MRI, particularly in a psychiatric setting.

In this proof-of-principle study we aimed to develop a simple neuroimaging protocol to differentiate between AD and geriatric depression which could form the basis of an optimized protocol for use in a clinical psychiatry context where MRI would typically be used as a first-line assessment for dementia and associated pathology. In this sense, optimized refers to the availability of clinical resources, not the biomarkers per se. We applied a two-step MRI driven approach exploiting the different degree of hippocampal volume loss present in both disorders to derive hippocampal volume thresholds for identifying participants who could first be diagnosed without a PET scan. Amyloid imaging using ^18^F-flutametamol was used to improve diagnostic accuracy only in cases where uncertainty remained. We compared the benefits of using this sequential model to the use of a single model incorporating both hippocampal volume and amyloid PET to classify participants.

## Methods

### Participants

Data from 27 participants with clinically probable AD* (*henceforth ‘AD’) and 41 age-matched currently depressed participants were included. The participants with clinically probable AD were recruited via 7 participating academic memory clinics as part of a General Electric (GE) Healthcare sponsored phase II clinical trial investigating ^18^F-flutemetamol imaging in AD and MCI (26). Subjects with geriatric depression were recruited from the Geriatric Psychiatry Department of one academic hospital as part of a larger study investigating cerebral amyloidosis in geriatric depression (23). The study was approved by the UZ Leuven Ethical Committee (S52151), and written informed consent was obtained from each subject in accordance with the Declaration of Helsinki.

### Participants with AD

Subjects older than 55 years were included. The AD diagnosis was based on the National Institute of Neurological and Communicative Diseases and Stroke– Alzheimer’s Disease and Related Disorders Association criteria for clinically probable AD and on the Diagnostic and Statistical Manual of Mental Disorders-IV criteria for dementia of Alzheimer type. The interval between the clinical diagnosis and study enrollment ranged between 1 month and 6 years. The Mini Mental State Examination (MMSE) score range for inclusion was 15 to 27. Eight AD subjects had a Clinical Dementia Rating scale (CDR) score of 0.5 and the remaining 19 a CDR of 1.

### Participants with geriatric depression

Subjects were included according to the following criteria: older than 60 years, having a major depressive disorder according to DSM IV criteria and not primarily referred for assessment of cognitive impairment.

Exclusion criteria for all participants were comorbid major psychiatric illness, previous or current alcohol and/or drug dependence, neurological illness (including stroke, transient ischemic attacks and dementia), medication precluding cognitive testing, metal implants precluding MRI scanning and ECT in the last 6 months prior to inclusion.

To ensure the samples were matched for mean age, and thus avoid the confounding effect of age as a diagnostic predictor in the logistic regression model, 7 subjects from the original n=48 De Winter et al cohort were subsequently excluded based solely on the criterion age >82 years.

### Imaging

#### Magnetic resonance imaging

Structural MRI data were acquired using an 8-channel head coil on a 3T Philips Intera scanner (Best, The Netherlands). High-resolution 3D turbo field echo (3DTFE) T1-weighted images were acquired with parameters: TR=9.6ms, TE=4.6ms, flip angle=8°, Slice thickness=1.2mm, in-plane voxel-size=0.98 x 0.98 x 1.2mm^3^, 182 axial slices.

#### Hippocampal volume

Two trained raters blinded to diagnosis manually delineated the hippocampus in native space. Manual editing was performed using ITK-SNAP version 2.4 (http://www.itksnap.org/pmwiki/pmwiki.php) in accordance with the HARMONIZED protocol guidelines (27). Hippocampal volumes were normalized based on total intracranial volume (TIV) using a validated method (28). TIV was obtained from an automated segmentation of grey matter, white matter and CSF (29). Inter-rater reliability was determined using randomly selected scans segmented at two time-points at least one month apart. The intra-class correlation coefficient (Cronbach’s α) was 0.95 for the left hippocampus and 0.98 for the right hippocampus. Inter-rater reliability was determined using 10 randomly selected scans from both subject groups. The intra-class correlation coefficient (Cronbach’s α) was 0.98 for the left hippocampus and 0.99 for the right hippocampus. Total normalised hippocampal volume was obtained by summing the left and right normalised hippocampal volumes.

#### 18F Flutemetamol PET imaging

18F Flutemetamol PET brain data of the AD cohort was acquired at 3 different scanning centers using a Biograph PET/CT scanner (Siemens, Erlangen, Germany), an ECAT EXACT HR+ scanner (Siemens), and a GE Advance (GE Healthcare, Milwaukee, US) scanner, respectively. The late life depression (LLD) cohort data were all obtained on the same Biograph PET/CT scanner (Siemens, Erlangen, Germany). The tracer was injected intravenously as a bolus in an antecubital vein (for the AD group: mean activity 161 MBq, SD= 25, range = 118-183; for the LLD group: mean activity, 149 MBq, SD=5, range=139–159). Image acquisition started 90 minutes after tracer injection and lasted for 30 minutes. Prior to the PET scan, a low-dose CT scan was performed for attenuation correction; for the HR+ scanner a Ge68 transmission scan was performed. Random and scatter corrections were also applied. Images were reconstructed to six frames of 5 minutes, using an ordered subsets expectation maximization algorithm (4 iterations x 16 subsets). All six PET frames were realigned to the first frame to correct for potential head motion. Subsequently, the realigned frames were summed to create one PET summed image. Individual T1-weighted MRI images were coregistered to this PET summed image. These MRI images were then normalized to Montreal Neurological Institute space, after which the normalization matrix was applied to the coregistered PET summed images. Standardized uptake value ratio (SUVR) images were calculated from the normalized PET summed image using cerebellar gray matter as reference region. The cerebellar gray matter volume of interest was derived from the Automated Anatomical Labeling Atlas (AAL) and was masked with the normalized subject specific segmented gray matter map to exclude white matter content.

#### Cerebral amyloid assessment

The mean SUVR value was calculated in five bilateral volumes of interest derived from the AAL atlas: lateral frontal, parietal, and temporal cortex and anterior and posterior cingulate cortex. A composite cortical SUVR value (SUVR_comp_) was calculated by averaging across these five volumes of interest. As with the cerebellar gray matter volume of interest, cortical volumes of interest were masked by the normalized subject-specific gray matter map (30).

### Statistical analysis

Two logistic regression model approaches were applied to test the relative benefit of using both hippocampal volume and amyloid PET measures in combination, or sequentially with HV first then amyloid, to classify subjects as having a diagnosis of depression or AD. In the first approach, a logistic regression model with both predictors is used. In the second approach, a logistic regression model with hippocampal volume as predictor is used in the first step and the the neuropathogically validated threshold of 1.38 for amyloid positivity based on 18F-Flutemetamol PET is used in the second step. The sensitivity and specificity to assign diagnosis were calculated for both approaches. P-values <0.05 were considered statistically significant. All analyses were conducted using SAS software version 9.4 (2015, Cary, USA).

#### Model 1: One step model

Logistic regression with both (total, normalised) hippocampal volume and SUVR_comp_ as predictors of AD classification.

#### Model 2: Sequential, two-step model

First, logistic regression with (total, normalised) hippocampal volume as a predictor of AD diagnosis was used to identify two hippocampal volume thresholds with an 85% probability to classify subjects as having respectively AD and LLD subjects. Based on these thresholds, patients were classified as AD, LLD or undetermined. Second, the subgroup of subjects unclassified in the first step were classified as AD or LLD based on the amyloid PET SUVR_comp_ threshold of 1.38 (AD >1.38 and LLD<=1.38). The threshold was derived from data from our centre for determining amyloid positivity(30).

## Results

### Population characteristics

The mean age of the depression group was 72.3 years (SD 6.6, range 59 -82 years) and the AD group was 69.6 (SD 7.0, range 60 -82 years). There were 29 females in the depression group (71%) and 15 (56%) in the AD group. The mean MMSE score was 24.4 (SD 3.8) for the depressed group and 23.3 (SD 2.2) for the AD group. There were no statistically significant differences in mean age (t(66)=1.62, p=0.11), mean MMSE score (t(45) = 1.37, p=0.16) or gender composition (χ^2^(1)=1.64, p=0.15) between groups. The mean total normalised hippocampal volume of the depressed and AD groups were 6500 mm^3^ (SD=694) and 5128 mm^3^ (SD=803) respectively. The median SUVR_comp_ of the depressed and AD groups were 1.33 (SD=0.19) and 1.97 (SD=0.30) respectively. Subjects with AD had a lower mean hippocampal volume (t(66)=7.49, p<0.001) and higher median amyloid load (χ^2^(1)=29.7, p<0.001) as compared to subjects with geriatric depression.

### Model 1 (One-step model)

The global null-hypothesis that all regression coefficients were zero in the logistic regression model including (total, normalised) hippocampal volume and SUVR_comp_ was rejected (Wald χ^2^(2) = 11.4, p=0.003). Within the model, both (total, normalised) hippocampal volume Wald χ^2^(1) = 6.46, p=0.011 and SUVR_comp_ (Wald χ^2^(1) = 11.0, p = 0.0009) were significant predictors of classification. Specifically, the lower the hippocampal volume and the higher the amount of amyloid, the higher the probability of having AD. Conversely, the higher the hippocampal volume and the lower the amount of amyloid, the higher the probability of depression. The sensitivity using both parameters to detect depression was 95%, and the specificity was 89%. In total 93 % of the subjects were correctly classified. Two subjects with AD (age: 74, 75) were incorrectly classified as having geriatric depression, whereas 3 subjects with depression (age: 74, 81, 82) were incorrectly classified as having AD.

#### Model 2 (Sequential, two-step model)

##### Step 1: Determination of diagnostic hippocampal volume thresholds

In the logistic regression model including (total, normalised) hippocampal volume only, hippocampal volume was a significant predictor of diagnosis, Wald χ^2^(1) = 18.3967, p <0.0001). Based on this model, hippocampal volume cut-off values of 4983 mm^3^ and 6393 mm^3^ were determined to classify subjects as having AD or depression respectively, with a probability of 0.85 (Figure 1). Using these cut-off values, 13 (19 %) of the subjects were correctly classified as having ‘AD’, 22 (32%) of the subjects were correctly classified as having depression, and 33 (49 %) of the subjects remained unclassified (being in-between both thresholds) (Figure 2). Note that by using this procedure, only one subject was incorrectly classified as having geriatric depression (see blue triangle at the upper right in figure 2).

**Figure 1.**
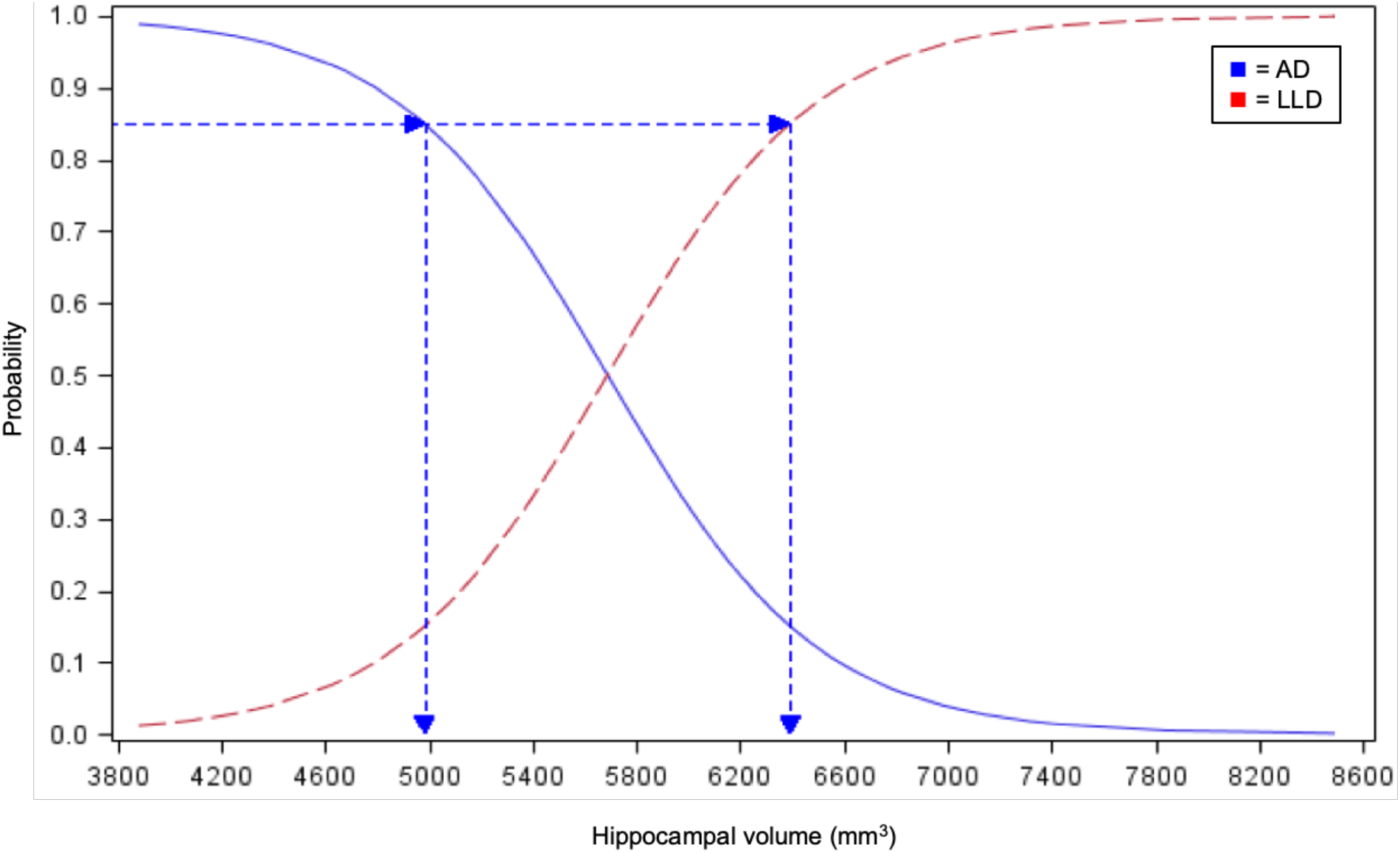
Probability of Alzheimer’s Disease or Late-life Depression diagnosis for a given normalised total hippocampal volume (mm^3^)

**Figure 2.**
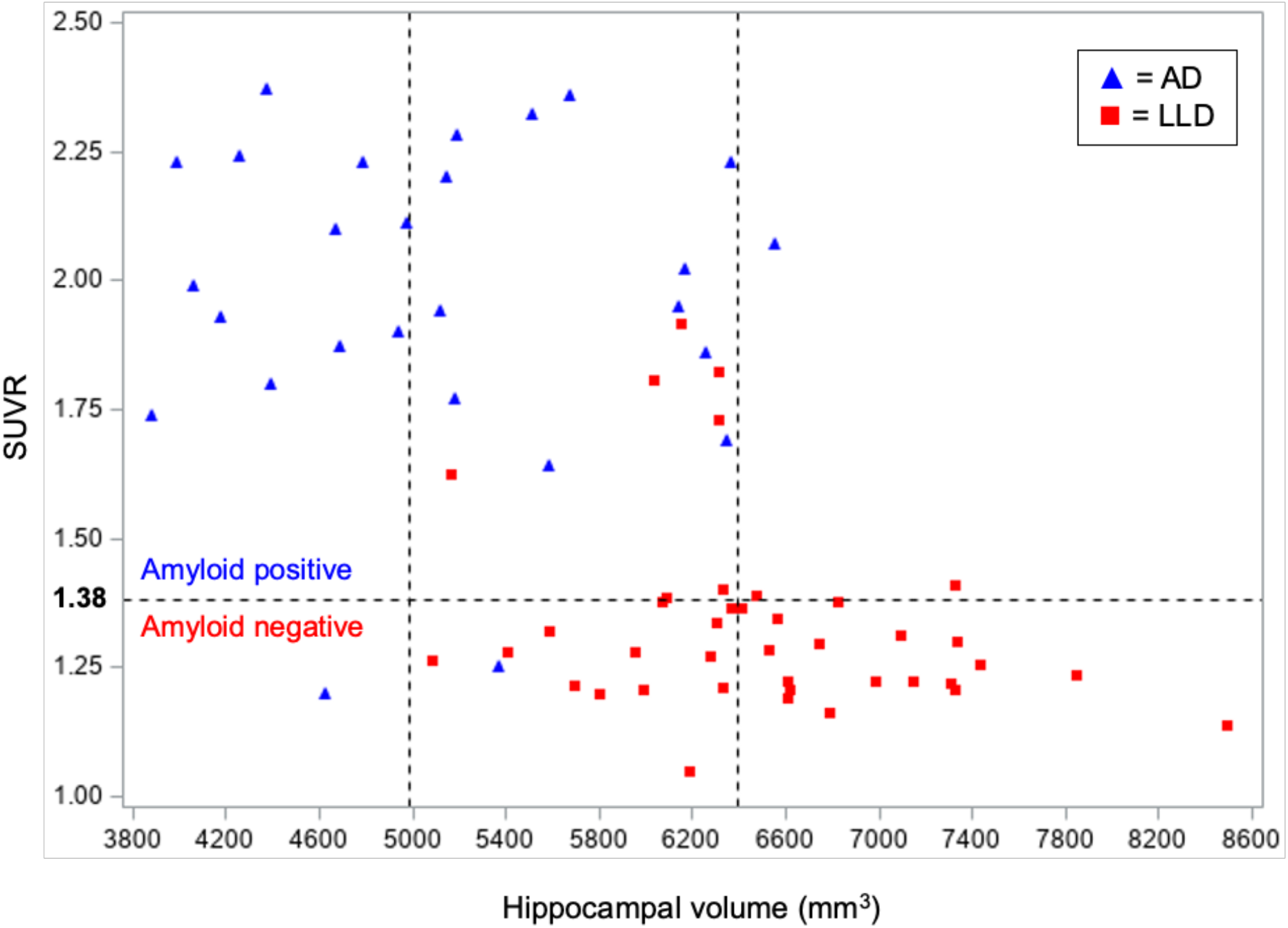
Classification of AD or Geriatric Depression diagnosis for a given amyloid load

>> Figure 1. Probability of Alzheimer’s Disease or Late-life Depression diagnosis for a given normalised total hippocampal volume (mm^3^) <<

##### Step 2: Using amyloid positivity to distinguish between AD and depression in unclassified subgroup

In this step subjects with an SUVR_comp_ below 1.38 were classified as having depression whereas subjects with a threshold above 1.38 were classified as having AD. These results can be appreciated visually in Figure 2. Based on the two-step procedure, the sensitivity to detect depression was 94% whereas the specificity was 78%. In total, 87% of the subjects were correctly classified according to their clinical diagnosis. Two subjects with AD (age: 66 and 75) were incorrectly classified as having LLD, whereas 7 subjects with LLD (age: 74, 75 76, 78, 81, 81, 82) were incorrectly classified as having AD.

>> Figure 2 Classification of AD or Geriatric Depression diagnosis for a given amyloid load <<

The results of both models are summarized in figure 3.

**Figure 3:**
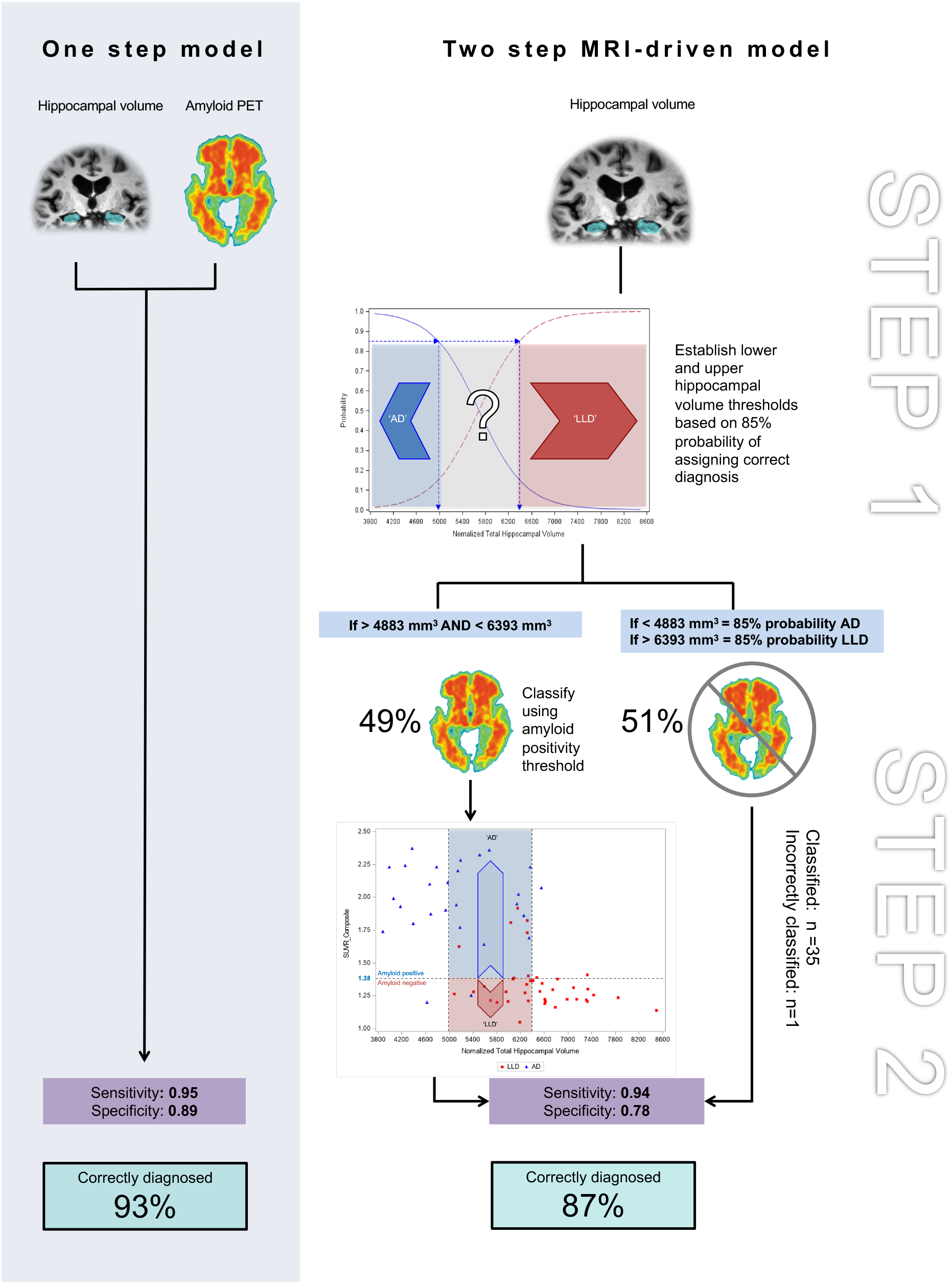
Summary of MRI / PET based protocol for improving the differential diagnosis of Geriatric Depression and Alzheimer’s Disease.

>> Figure 3: Summary of MRI / PET based protocol for improving the differential diagnosis of Geriatric Depression and Alzheimer’s Disease. <<

## Discussion

The main objective of this proof of concept study was to investigate the use of two imaging biomarkers, hippocampal volume and amyloid PET, to improve the differential diagnosis of/classification between AD and geriatric depression in a clinical psychiatry setting. Furthermore, we aimed to compare the relative benefits of using a sequential model to a single model that prioritizes hippocampal volume given that MRI is more commonly used than amyloid PET as a first line assessment in psychiatric practice.

Both hippocampal volume and amyloid load proved to be significant predictors of diagnosis, with lower hippocampal volume and higher amyloid load increasing the probability of having AD, and higher volume and lower amyloid load increasing the probability of depression. Given the challenges differentiating between geriatric depression and AD, the use of MRI-based hippocampal volume and amyloid PET could significantly decrease the number of people incorrectly diagnosed.

Previous studies using hippocampal volume to distinguish between AD and LLD led to diverging conclusions. Joko et al.(31) found an overall trend of corrected hippocampal volumes with the smallest volumes found in AD subjects, then aMCI, MDD and finally normal controls. These findings are in line with ours, where hippocampal volumes below 4883 mm^3^ indicated AD and volumes above 6393 mm^3^ indicated LLD. Besides differences in corrected hippocampal volume, Joko et al (31) also found regional differences between subject groups. Overall atrophy was seen in AD and aMCI whereas in MDD atrophy was more localized in the anterior hippocampal formation compared to controls. According to the authors, these regional differences could be used as a tool for differentiating between AD, aMCI and MDD.

Similarly, O’Brien et al (32) found that the degree of anterior hippocampal atrophy could be used in differentiating AD from MDD. Hippocampal atrophy was assessed using a 4-point scale, anterior atrophy below a score of 2 was more suggestive for MDD whereas a score above 2 was more suggestive for AD with a sensitivity of 93%, a specificity of 84%. Overall 89% of cases were correctly diagnosed. The discrepancy in classification rate with ours based only on hippocampal volume (51%) is interesting. O’Brien et al used low resolution 5mm 2D T1-weighted scans from a 0.3T scanner and a visual rating scale of temporal lobe atrophy in contrast to our higher resolution quantitative analysis which should theoretically yield more accurate results. Future work focusing on the anterior hippocampus may therefore improve our classification further. A lateralization effect in hippocampal volume decrease was also reported by Sahin et al with the left hippocampal volume being significantly reduced in the AD group compared to the depression group (33). One pilot study (34) focused on hippocampal and entorhinal cortex (EC) volume reduction (assessed by manual segmentation) between 6 LLD and 12 aMCI subjects with and without depressive symptoms (not meeting DSM IV criteria for MDD). The aMCI subjects with depressive symptoms could be categorized as subjects with MBI and concurrent MCI. Hippocampal volume reduction was larger in aMCI compared to aMCI with depressive symptoms and geriatric depression, however the volume differences between groups did not reach statistical significance. The authors propose that people with aMCI and depressive symptoms are closer to geriatric depression than aMCI and thus should be considered as a different entity. A meta-analysis of Boccia et al (35) compared grey matter changes in AD and LLD. As expected, both were linked to a reduction of bilateral hippocampal volume but only AD was correlated with greater atrophy in the left anterior hippocampus and bilateral posterior cingulate cortex (PCC). Note that caution is needed when comparing the results of these studies due to the variation in assessment of hippocampal volume. For this reason, standard operating procedures for MR-based manual hippocampal segmentation were recently developed which were also applied in our study (27).

In the second step of the two-step model, we used amyloid PET in the remaining 49% of subjects who had a (total, normalised) hippocampal volume between 4883 mm^3^ and 6393 mm^3^. To our knowledge, there are no previous studies that have investigated the use amyloid PET as a tool in differentiating AD from geriatric depression, although the technique is recognized as an important tool to improve diagnostic certainty in subjects with possible AD(36). In contemporary psychiatric practice, the accessibility of amyloid scans is more limited, therefore the sequential two-step model presented in this study accounts for this by using HV measurements as a gatekeeper in case amyloid-PET is not available. Using this approach, we could negate the need for an amyloid PET exam in 51% of subjects. Nevertheless, it should be noted that when using only amyloid PET in a logistic regression model, 81% of cases would be correctly classified, making it a more useful biomarker than HV for discrimination in situations where amyloid PET is more accessible than MRI.

Several limitations of this study need to be addressed. First, the small sample size and retrospective nature limits generalization of the results. Second, the reported thresholds to differentiate AD from LLD using hippocampal volume are based on the sample itself. Therefore, these cut off values are merely indicative and cannot be used as independent thresholds. Third, based on the inherent limitations of a clinical diagnosis of LLD we cannot rule out that some of these subjects were in fact not (preclinical) AD subjects with depression. Furthermore, the use of technical biomarkers might give the impression that a categorical distinction between AD and LLD is always possible. However, comorbid pathology cannot be ruled out based on this procedure. Fourth, we did not take into account variables that could influence hippocampal volume directly or indirectly such as treatment with antidepressants or vascular brain changes. Fifth, manual assessment of hippocampal volume limits its applicability to daily clinical practice. Sixth, this study focused only on amyloid PET, however the FDG PET may also be useful to investigate given its more widespread availability. Lastly, we did not include prodromal AD subjects. In future studies it would be interesting to compare hippocampal volume changes and amyloid deposition in MCI and MBI subjects in comparison to AD and geriatric depression subjects.

The current results suggest that it may be advantageous to use a sequential MRI driven clinical protocol instead of a combined approach in a psychiatry setting given only 49% of the sample had to be tested with amyloid PET in order to be diagnosed correctly, and only 6% fewer subjects were incorrectly classified compared to the combined model. Due to the small sample size and variability in current hippocampal volume measurement techniques (manual vs automated) the precise thresholds calculated in this study are only indicative. However, with the advent and regulation of quantitative MRI approaches within radiology, more accessible and standardized hippocampal volume and other structural MRI measures are increasingly becoming available (37).

In conclusion, the proposed two-step model forms the basis of an efficient clinical imaging protocol to differentiate AD and geriatric depression, and should be cross-validated in a new, larger, sample using standardized, clinically accessible, automated hippocampal volume measures across the depression-dementia spectrum.

## Data Availability

The data that support the findings of this study are available on reasonable request from the corresponding author

## Acknowledgements

The authors would like to thank Gill Farrer and Paul Sherwin for their feedback on the manuscript.

## Funding

This study was supported by Research Foundation Flanders (FWO) project G074609, and Research Grant Old Age Psychiatry UZ Leuven R94859. The tracer [18F]-flutemetamol was provided free of charge by GE for this investigator-led analysis. Additionally, MV, FB, LE have received funding from the FWO (G0C0319N), and along with JVdS are supported by KU Leuven: C24/18/095 and The Sequoia Fund. RV has received funding from the Foundation for Alzheimer Research (SAO-FRMA) (09013, 11020, 13007) and KU Leuven OT/08/056, OT/12/097.

## Declaration of Conflicting Interests

RV has a consultancy agreement from AC Immune, and his institution has a clinical trial agreement with AbbVie and Roche. K. Van Laere has received contract research grants through KU Leuven from Merck, Janssen Pharmaceuticals, Abide, UCD, Cerveau, Syndesi, Eikonizo, GE Healthcare, and Curasen; he has received speaker fees from GE Healthcare. Part of the data used in this study was derived from a GE-sponsored phase II clinical trial. The sponsors had no role in the study design; collection, analysis, or interpretation of data; nor imposed any restrictions regarding the submission of this report for publication.

## Notes

### Author Declarations

UZ Leuven Ethics Committee (S52151)

## References

1. Robbins TW, Elliott R, Sahakian BJ. Neuropsychology--dementia and affective disorders. Br Med Bull. 1996;52(3):627–43.

2. Yesavage J. Differential diagnosis between depression and dementia. Am J Med. 1993;94(5A):23S–8S.

3. Downing LJ, Caprio TV, Lyness JM. Geriatric psychiatry review: differential diagnosis and treatment of the 3 D’s - delirium, dementia, and depression. Curr Psychiatry Rep. 2013;15(6):365.

4. Visser PJ, Verhey FR, Ponds RW, Kester A, Jolles J. Distinction between preclinical Alzheimer’s disease and depression. J Am Geriatr Soc. 2000;48(5):479–84.

5. Zubenko GS, Zubenko WN, McPherson S, Spoor E, Marin DB, Farlow MR, et al. A collaborative study of the emergence and clinical features of the major depressive syndrome of Alzheimer’s disease. Am J Psychiatry. 2003;160(5):857–66.

6. Lyketsos CG, Olin J. Depression in Alzheimer’s disease: overview and treatment. Biol Psychiatry. 2002;52(3):243–52.

7. Ownby RL, Crocco E, Acevedo A, John V, Loewenstein D. Depression and risk for Alzheimer disease: systematic review, meta-analysis, and metaregression analysis. Arch Gen Psychiatry. 2006;63(5):530–8.

8. Jorm AF. History of depression as a risk factor for dementia: an updated review. Aust N Z J Psychiatry. 2001;35(6):776–81.

9. Sheline YI, Wang PW, Gado MH, Csernansky JG, Vannier MW. Hippocampal atrophy in recurrent major depression. Proc Natl Acad Sci U S A. 1996;93(9):3908–13.

10. Bell-McGinty S, Butters MA, Meltzer CC, Greer PJ, Reynolds CF, Becker JT. Brain morphometric abnormalities in geriatric depression: long-term neurobiological effects of illness duration. Am J Psychiatry. 2002;159(8):1424–7.

11. Panza F, Frisardi V, Capurso C, D’Introno A, Colacicco AM, Imbimbo BP, et al. Late-life depression, mild cognitive impairment, and dementia: possible continuum? Am J Geriatr Psychiatry. 2010;18(2):98–116.

12. Singh-Manoux A, Dugravot A, Fournier A, Abell J, Ebmeier K, Kivimäki M, et al. Trajectories of Depressive Symptoms Before Diagnosis of Dementia: A 28-Year Follow-up Study. JAMA Psychiatry. 2017;74(7):712–8.

13. Ismail Z, Smith EE, Geda Y, Sultzer D, Brodaty H, Smith G, et al. Neuropsychiatric symptoms as early manifestations of emergent dementia: Provisional diagnostic criteria for mild behavioral impairment. Alzheimers Dement. 2016;12(2):195–202.

14. Cieslak A, Smith EE, Lysack J, Ismail Z. Case series of mild behavioral impairment: toward an understanding of the early stages of neurodegenerative diseases affecting behavior and cognition. Int Psychogeriatr. 2018;30(2):273–80.

15. Palmer K, Di Iulio F, Varsi AE, Gianni W, Sancesario G, Caltagirone C, et al. Neuropsychiatric predictors of progression from amnestic-mild cognitive impairment to Alzheimer’s disease: the role of depression and apathy. J Alzheimers Dis. 2010;20(1):175–83.

16. Gasser AI, Salamin V, Zumbach S. [Late life depression or prodromal Alzheimer’s disease: Which tools for the differential diagnosis?]. Encephale. 2018;44(1):52–8.

17. Albert MS, DeKosky ST, Dickson D, Dubois B, Feldman HH, Fox NC, et al. The diagnosis of mild cognitive impairment due to Alzheimer’s disease: recommendations from the National Institute on Aging-Alzheimer’s Association workgroups on diagnostic guidelines for Alzheimer’s disease. Alzheimers Dement. 2011;7(3):270–9.

18. Dubois B, Feldman HH, Jacova C, Dekosky ST, Barberger-Gateau P, Cummings J, et al. Research criteria for the diagnosis of Alzheimer’s disease: revising the NINCDS-ADRDA criteria. Lancet Neurol. 2007;6(8):734–46.

19. Jack CR, Albert MS, Knopman DS, McKhann GM, Sperling RA, Carrillo MC, et al. Introduction to the recommendations from the National Institute on Aging-Alzheimer’s Association workgroups on diagnostic guidelines for Alzheimer’s disease. Alzheimers Dement. 2011;7(3):257–62.

20. Geerlings MI, Gerritsen L. Late-Life Depression, Hippocampal Volumes, and Hypothalamic-Pituitary-Adrenal Axis Regulation: A Systematic Review and Meta-analysis. Biol Psychiatry. 2017;82(5):339–50.

21. Gerritsen L, Comijs HC, van der Graaf Y, Knoops AJ, Penninx BW, Geerlings MI. Depression, hypothalamic pituitary adrenal axis, and hippocampal and entorhinal cortex volumes--the SMART Medea study. Biol Psychiatry. 2011;70(4):373–80.

22. Taylor WD, McQuoid DR, Payne ME, Zannas AS, MacFall JR, Steffens DC. Hippocampus atrophy and the longitudinal course of late-life depression. Am J Geriatr Psychiatry. 2014;22(12):1504–12.

23. De Winter FL, Emsell L, Bouckaert F, Claes L, Jain S, Farrar G, et al. No Association of Lower Hippocampal Volume With Alzheimer’s Disease Pathology in Late-Life Depression. Am J Psychiatry. 2017;174(3):237–45.

24. Jansen WJ, Ossenkoppele R, Knol DL, Tijms BM, Scheltens P, Verhey FR, et al. Prevalence of cerebral amyloid pathology in persons without dementia: a meta-analysis. JAMA. 2015;313(19):1924–38.

25. Johnson KA, Minoshima S, Bohnen NI, Donohoe KJ, Foster NL, Herscovitch P, et al. Update on appropriate use criteria for amyloid PET imaging: dementia experts, mild cognitive impairment, and education. Amyloid Imaging Task Force of the Alzheimer’s Association and Society for Nuclear Medicine and Molecular Imaging. Alzheimers Dement. 2013;9(4):e106–9.

26. Vandenberghe R, Van Laere K, Ivanoiu A, Salmon E, Bastin C, Triau E, et al. 18F-flutemetamol amyloid imaging in Alzheimer disease and mild cognitive impairment: a phase 2 trial. Ann Neurol. 2010;68(3):319–29.

27. Boccardi M, Bocchetta M, Apostolova LG, Barnes J, Bartzokis G, Corbetta G, et al. Delphi definition of the EADC-ADNI Harmonized Protocol for hippocampal segmentation on magnetic resonance. Alzheimers Dement. 2015;11(2):126–38.

28. Jack CR, Twomey CK, Zinsmeister AR, Sharbrough FW, Petersen RC, Cascino GD. Anterior temporal lobes and hippocampal formations: normative volumetric measurements from MR images in young adults. Radiology. 1989;172(2):549–54.

29. Jain S, Sima DM, Ribbens A, Cambron M, Maertens A, Van Hecke W, et al. Automatic segmentation and volumetry of multiple sclerosis brain lesions from MR images. Neuroimage Clin. 2015;8:367–75.

30. Adamczuk K, Schaeverbeke J, Nelissen N, Neyens V, Vandenbulcke M, Goffin K, et al. Amyloid imaging in cognitively normal older adults: comparison between (18)F-flutemetamol and (11)C-Pittsburgh compound B. Eur J Nucl Med Mol Imaging. 2016;43(1):142–51.

31. Joko T, Washizuka S, Sasayama D, Inuzuka S, Ogihara T, Yasaki T, et al. Patterns of hippocampal atrophy differ among Alzheimer’s disease, amnestic mild cognitive impairment, and late-life depression. Psychogeriatrics. 2016;16(6):355–61.

32. O’Brien JT, Desmond P, Ames D, Schweitzer I, Chiu E, Tress B. Temporal lobe magnetic resonance imaging can differentiate Alzheimer’s disease from normal ageing, depression, vascular dementia and other causes of cognitive impairment. Psychol Med. 1997;27(6):1267–75.

33. Sahin S, Okluoglu Önal T, Cinar N, Bozdemir M, Çubuk R, Karsidag S. Distinguishing Depressive Pseudodementia from Alzheimer Disease: A Comparative Study of Hippocampal Volumetry and Cognitive Tests. Dement Geriatr Cogn Dis Extra. 2017;7(2):230–9.

34. Morin JF, Mouiha A, Pietrantonio S, Duchesne S, Hudon C. Structural neuroimaging of concomitant depressive symptoms in amnestic mild cognitive impairment: a pilot study. Dement Geriatr Cogn Dis Extra. 2012;2(1):573–88.

35. Boccia M, Acierno M, Piccardi L. Neuroanatomy of Alzheimer’s Disease and Late-Life Depression: A Coordinate-Based Meta-Analysis of MRI Studies. J Alzheimers Dis. 2015;46(4):963–70.

36. Rice L, Bisdas S. The diagnostic value of FDG and amyloid PET in Alzheimer’s disease-A systematic review. Eur J Radiol. 2017;94:16–24.

37. Struyfs H, Sima DM, Wittens M, Ribbens A, Pedrosa de Barros N, Phan TV, et al. Automated MRI volumetry as a diagnostic tool for Alzheimer’s disease: Validation of icobrain dm. Neuroimage Clin. 2020;26:102243.

